# Pharmacodynamic effects of semorinemab on plasma and CSF biomarkers of Alzheimer’s disease pathophysiology

**DOI:** 10.1101/2024.04.18.24306056

**Authors:** Stephen Schauer, Balazs Toth, Julie Lee, Lee A. Honigberg, Vidya Ramakrishnan, Jenny Jiang, Gwendlyn Kollmorgen, Anna Bayfield, Norbert Wild, Jennifer Hoffman, Ryan Ceniceros, Michael Dolton, Sandra Sanabria Bohorquez, Casper C. Hoogenraad, Kristin R. Wildsmith, Edmond Teng, Cecilia Monteiro, Veronica Anania, Felix L. Yeh

## Abstract

**INTRODUCTION:** Semorinemab, an anti-tau monoclonal antibody, was evaluated in two Phase II trials as a disease-modifying treatment for Alzheimer’s disease (AD). Plasma and cerebrospinal fluid (CSF) samples were collected from trial participants to evaluate the pharmacodynamic effects of semorinemab and elucidate its mechanism of action.

**METHODS:** Qualified immunoassays were used to measure plasma and CSF biomarkers of tau, amyloidosis, glial activity, neuroinflammation, synaptic function, and neurodegeneration from participants enrolled in the Tauriel (NCT03289143) and Lauriet (NCT03828747) Phase II trials in prodromal-to-mild (P2M) and mild-to-moderate (M2M) AD.

**RESULTS:** Significant increases in plasma phosphorylated Tau 181 (pTau181) and CSF Chitinase-3-like protein 1 (YKL-40) followed administration of semorinemab in both studies. In the Lauriet study, plasma glial fibrillary protein (GFAP) levels rose progressively over the study period in the placebo group, but remained stable over time with the administration of semorinemab. In contrast, this was not observed in the Tauriel study. Semorinemab had no consistent impact on other biomarkers of AD pathophysiology that were evaluated.

**DISCUSSION:** Semorinemab engages and stabilizes plasma pTau181 in a manner consistent with previously reported data [1,2], and levels do not decrease after prolonged drug exposure. Increases to CSF YKL-40 suggest that semorinemab may stimulate microglia activation, while stabilization of plasma GFAP levels in Lauriet participants indicate that semorinemab may moderate reactive gliosis in M2M AD.

## BACKGROUND

Alzheimer’s disease is the most common form of dementia, representing 60-80% of cases. It is progressive, with no known cure, and an estimated 6.7 million people in the United States are afflicted with the disease [3]. AD is characterized by two main pathological hallmarks: extracellular amyloid plaques composed of aggregated amyloid-beta (Aβ) peptides, and intracellular neurofibrillary tangles that consist of the protein tau [4]. The spatial temporal burden of cortical tau neurofibrillary tangles tracks more closely with neuronal loss and antemortem cognitive measures than amyloid [5–8] suggesting, along with mechanistic data from *in vitro* and *in vivo* models [9], that tau is an active contributor to AD pathogenesis and a viable therapeutic target. Tau pathology spreads in a predictable rostral-caudal sequence in AD [5,10] implying that it may be released extracellularly and propagates across interconnected brain networks in a prion-like fashion [11]. Semorinemab, a humanized anti-tau IgG4 monoclonal antibody under development for AD, is hypothesized to target the proliferation and accumulation of tau pathology by intercepting and clearing tau from the extracellular space.

Two Phase II studies in different AD patient populations were conducted: Tauriel (NCT03289143), a Phase II study of semorinemab in prodromal-to-mild (P2M) AD patients, and Lauriet (NCT03828747), a Phase II study in mild-to-moderate (M2M) AD patients. In Tauriel, semorinemab failed to demonstrate clinical efficacy at doses of 1500, 4500, and 8100 mg for 73 weeks. In contrast, a 4500 mg dose of semorinemab for 49 or 61 weeks was associated with a significant slowing in progression (versus placebo) in a co-primary endpoint of cognition (Alzheimer’s Disease Assessment Scale-Cognitive Subscale 11 (ADAS-Cog11)) but it was not accompanied by improvements in daily function (Alzheimer’s Disease Cooperative Study-Activities of Daily Living (ADCS-ADL)). In an attempt to reconcile conflicting clinical outcomes with evidence of CSF tau target engagement in both studies [1,2], exploratory post-hoc assessments of AD pathophysiology biomarkers were performed to gain clearer insight into semorinemab’s mechanism of action.

## METHODS

### P2M and M2M AD participants from Tauriel and Lauriet

In Tauriel (NCT03289143), patients with prodromal or mild AD were recruited per criteria from the National Institute on Aging and Alzheimer’s Association (NIA-AA). Participants were aged 50-80 years old, with Mini-Mental State Examination (MMSE) scores between 20-30, Clinical Dementia Rating (CDR) Global Score of 0.5 or 1, episodic memory deficit by Repeatable Battery for the Assessment of Neuropsychological Status Delayed Memory Index (RBANS DMI) ≤ 85, and positive for cerebral amyloid pathology by an amyloid-beta (Aβ) positron emission tomography (PET) visual read or by reduced CSF amyloid-β (1-42) concentrations. Patients were randomized 2:3:2:3, and received intravenous (IV) doses of 1500, 4500, or 8100 mg semorinemab or placebo administered every two weeks (Q2W) for doses 1 through 3, and then every four weeks (Q4W) until the end of the DBL study period (Week 73) [1]. In Lauriet (NCT03828747), 272 patients with mild-to-moderate AD were recruited; participants were aged 50-85, with MMSE scores between 16-21, CDR-GS Global Score of 1 or 2, and positive for amyloidosis by positron emission tomography (PET) imaging or reduced CSF amyloid-β (1-42) concentrations. Participants were randomized 1:1 to receive either placebo or 4500 mg Semo IV Q2W for doses 1 through 3, and then Q4W (every month) until the end of the DBL period (Cohort 1, Week 49). However, due to the COVID-19 pandemic, a subset of patients (n=76) missed more than two doses during the double-blind treatment period. To account for the potential effect of missed doses, the double blind period was extended for an additional 12 weeks for these patients (Cohort 2, Week 61). Serum pharmacokinetics demonstrated that these patients had the same overall exposure as patients who missed none or 1 dose. Patients who completed the double-blind treatment period had the option to enter into an open-label extension, during which all patients received semorinemab 4500 mg Q4W for 96 weeks. Patients provided plasma samples for pharmacodynamic and pharmacokinetic analyses at baseline (pre-dose) and Weeks 5, 25, 49 or 61. CSF collection was optional; of 272 patients, 48% (n=131) provided CSF at baseline and 19% (n=53) provided CSF at follow-up. Identical serum pharmacokinetics between the Week 49 and 61 cohorts enabled pooling of CSF results from each cohort to increase the statistical power [2]. Exploratory CSF and plasma analyses were restricted exclusively to samples from participants who provided informed consent. Baseline demographics of the study populations are detailed in **Table 1**.

**Table 1:** Demographics of P2M and M2M participants at baseline and clinical information.

|  | Tauriel | Lauriet |
| --- | --- | --- |
| Study Identifier | NCT03289143 | NCT03828747 |
| Diagnosis | Prodromal-to-Mild AD | Mild-to-Moderate AD |
| n | 431 | 267 |
| Age (mean (SD)) | 69.3 (7.09) | 71.8 (8.02) |
| Sex, M/F (n,%) | F: 241 (55.9)<br>M: 190 (44.1) | F: 173 (64.8)<br>M: 94 (35.2) |
| APOE4 +/- (n, %) | APOE4+: 316 (73.3)<br>APOE4-: 115 (26.7) | APOE4+: 167 (62.5)<br>APOE4-: 100 (37.5) |
| MMSE (mean (SD)) | 23.3 (2.68) | 18.2 (2.04) |

### Biomarkers

Exploratory fluid biomarker analyses of participant CSF and plasma samples were conducted to examine various aspects of AD pathophysiology, including measures of tau, glial activity, neuroinflammation, synaptic integrity, neurodegeneration, and amyloidosis. A detailed list of all biomarkers, matrices, and the studies in which they were measured can be found in **Table 2**.

**Table 2:** Summary of biomarker analyses.

| Biomarker | Matrices | Biology | Studies | Platform |
| --- | --- | --- | --- | --- |
| Total tau | CSF, Plasma | Tau burden, target engagement | Tauriel, Lauriet | NeuroToolKit |
| N-terminal tau | CSF | Target engagement | Tauriel, Lauriet | IPMS |
| pTau181 | CSF, Plasma | Tau burden, target engagement | Tauriel, Lauriet | CE-marked Elecsys (CSF), Elecsys RPA (plasma) |
| pTau217 | CSF, Plasma | Tau burden, target engagement | Tauriel, Lauriet (Plasma only) | Simoa (CSF), Elecsys RPA (Plasma) |
| CXCL10 | CSF | Glial activity, neuroinflammation | Tauriel, Lauriet | Simple Plex |
| GFAP | CSF, Plasma | Glial activity, neuroinflammation | Tauriel, Lauriet | NeuroToolKit |
| S100b | CSF | Glial activity, neuroinflammation | Tauriel, Lauriet | NeuroToolKit |
| sTREM2 | CSF | Glial activity, neuroinflammation | Tauriel, Lauriet | NeuroToolKit |
| YKL-40 | CSF, Plasma | Glial and lysosomal activity | Tauriel (CSF only), Lauriet | NeuroToolKit |
| Neurogranin | CSF | Synaptic integrity and function | Tauriel, Lauriet | NeuroToolKit |
| SNAP-25 | CSF | Synaptic integrity and function | Tauriel, Lauriet | NeuroToolKit |
| Neuronal pentraxin-2 | CSF | Synaptic integrity and function | Tauriel, Lauriet | NeuroToolKit |
| Alpha-synuclein | CSF | Synaptic integrity and function | Tauriel, Lauriet | NeuroToolKit |
| IL-6 | CSF | Neuroinflammation | Tauriel, Lauriet | NeuroToolKit |
| Lipocalin | CSF | Neuroinflammation | Tauriel, Lauriet | Simple Plex |
| β-amyloid (1-42) | CSF | Amyloidosis | Tauriel, Lauriet | CE-marked Elecsys β-Amyloid (1-42) |
| β-amyloid (1-40) | CSF | Amyloidosis | Tauriel, Lauriet | NeuroToolKit |
| Neurofilament-Light | CSF, Plasma | Neurodegeneration | Tauriel, Lauriet | NeuroToolKit |
| Neurofilament-Heavy | CSF | Neurodegeneration | Tauriel, Lauriet | Simple Plex |

### Elecsys immunoassay measurements of CSF and plasma samples

ElecsysⓇ electrochemiluminescence immunoassays were used to measure CSF and plasma samples. Plasma pTau181 was measured blinded on a cobasⓇ e 402 analyzer using a Robust Prototype Assay (RPA) with a special extended measurement range. Plasma pTau217 was measured blinded using an RPA on a cobas e 411 instrument. A suite of Elecsys assays collectively known as the Roche NeuroToolKit (NTK), which include both RPAs and CE-marked in vitro diagnostic assays, was used to measure concentrations of β-amyloid 1-42 (Aβ1-42), β-amyloid 1-40 (Aβ1-40), α-synuclein, glial fibrillary acidic protein (GFAP), interleukin-6 (IL-6), neurogranin (Ng), neurofilament light chain (NfL), neuronal pentraxin-2 (NPTX2), synaptosomal-associated protein 25kDa (SNAP-25), S100 calcium-binding protein B (S100B), soluble triggering receptor expressed on myeloid cells 2 (sTREM2), total tau (tTau), phosphorylated tau T181 (pTau181), and YKL-40 in the CSF, and GFAP, NfL, tTau, and YKL-40 in plasma (**Table 2**). These assays were validated to Clinical and Laboratory Standards Institute (CLSI) standards and demonstrated high intra- and inter-assay precision, which enabled singlicate determinations. Blinded measurements were performed on cobasⓇ e 411 and e 601 instruments (Roche Diagnostics International Ltd, Rotkreuz, Switzerland) at Covance CLS sites in Geneva (samples originating in Europe) and Indianapolis, IA (sample originating in the United States). Concentrations were calculated from 2-point calibration curves, and two recombinant protein quality controls were required to quantify within 21% of their established concentration for data to be acceptable.

### Simple Plex immunoassay measurements of CSF samples

C-X-C motif chemokine ligand 10 (CXCL10), lipocalin (LCN), neurofilament heavy Chain (NfH) from CSF were measured with SimplePlex assays on the ProteinSimple Ella^TM^ platform (Bio-Techne, San Jose, CA). Baseline and follow-up samples were measured in pairs in a single batch. Concentrations were calculated from factory calibration curves specific to each assay lot, and the intra-assay coefficient of variation (CV) was required to be ≤20% for data acceptability.

### Simoa immunoassay measurements of CSF samples

Concentrations of CSF pTau217 were measured with a custom single molecule array (Simoa) assay (Quanterix, Billerica, MA) developed in-house. The analyte was captured with a monoclonal antibody specific for the pTau217 epitope (Roche Diagnostics, GmbH, Penzberg, Germany), and then detected with biotinylated monoclonal antibody specific for the projection domain of tau (125B11H3, Genentech Inc., South San Francisco, CA). Samples were analyzed on the HD-1 system (Quanterix, Billerica, MA) where fluorescent signal in measured samples was converted to an average number of enzymes per bead (AEB). Concentrations were interpolated from a recombinant pTau calibration curve. Samples were analyzed in duplicate, with baseline and follow-up measured in the same assay run. Recombinant protein quality controls were required to quantify within 20% of their established concentration for data acceptability.

### IPMS for N-term tau

CSF N-terminal tau concentrations were measured by targeted LC-MS analysis. Protein A cartridges were crosslinked with an anti-N-terminal tau antibody (Ab62, Genentech, Inc.) on the AssayMAP Bravo Platform (Agilent, Santa Clara, CA). Patient CSF samples were randomized, and all available patient timepoints were measured within an assay run. CSF was clarified with perchloric acid and a centrifugation step, denatured, and then digested with lysyl-endopeptidase overnight. The peptides derived from this step were oxidized, purified on RP-S cartridges (Agilent, Santa Clara, CA), eluated, and then spiked with a stable isotope peptide standard. The N-terminal tau peptides were then captured with the Ab-62 cross-linked cartridges and eluted with 1% formic acid. The samples were injected into an M5 MicroLC (Sciex, Redwood City, CA) coupled to a 6500 QTRAP mass spectrometer (Sciex) operated in MRM mode. Four transition ions (y14, y15, y13, y9) for the precursor ion (A[+42]EPRQEFEVM[+16]EDHAGTYGLGDRK, 4+) were monitored for quantification. Raw data files were processed with Skyline software (University of Washington, USA), and concentrations were determined by using the normalized peak area of the endogenous peptide to the peak area of the stable isotope peptide standard against 8-point calibration curves with a range of 0.78-200 fmol/mL. Each 96 well plate of samples included two replicates of a pooled QC CSF lot and calibration curve. At least one QC replicate was required to quantify within 20% of the N-terminal tau established concentration for data acceptability.

### Statistical analysis

Comparisons between baseline biomarker concentrations in Tauriel and Lauriet were assessed for significance with a Mann-Whitney U test. P-values for comparisons between treatment groups within each study were calculated using a Student’s *t*-test. Spearman’s rho values were calculated to represent the strength of the correlation between biomarkers and clinical scores at baseline. Unless otherwise noted as nominally significant, p-values were adjusted for multiple comparisons using the False Discovery Rate (FDR) method, with the alpha set to 0.05 as a determination of significance. When conducting aggregated analyses of biomarkers across Weeks 49 and 61 in Lauriet, the data was annualized prior to analysis. Statistical assessments were performed with SAS statistical software version 9.4 (SAS Institute), R software version 4.3.3 (R Foundation), and Prism version 8.4.1 (GraphPad Software).

## RESULTS

### GFAP

Plasma and CSF GFAP levels were quantified in Lauriet and Tauriel participants to determine if semorinemab treatment was associated with changes in glial activity.

In each trial, pre-dose baseline plasma GFAP levels were comparable across the placebo and treatment groups (**Figure 1A**). In Lauriet, placebo plasma GFAP levels (expressed as the median percent change from pre-dose baseline levels ± standard error) increased as the study progressed (Week 49: 9.52 ± 2.56%, Week 61: 12.5 ± 5.96%) (**Figure 1B**). In contrast, plasma GFAP levels stabilized near baseline levels following administration of 4500 mg semorinemab (Week 49: 4.48 ± 2.42%, Week 61: 1.89 ± 4.23%). The difference between the baseline normalized changes in the placebo and 4500 mg semorinemab groups at Weeks 49 and 61 was not significant (**Figure 1B)**. An examination of unadjusted plasma GFAP concentrations was consistent with these results. The median (± standard error) post-dose concentration of plasma GFAP in each treatment cohort increased in parallel until Week 25, and then separated at Weeks 49 and 61. Plasma GFAP levels stabilized in the 4500 mg semorinemab group while concentrations in the placebo group continued to accumulate for the duration of the study. In contrast to the baseline-normalized percent change analysis, the difference between plasma GFAP concentrations in the 4500 mg semorinemab group (166 ± 14.0 pg/mL) and placebo group (191 ± 25.4 pg/mL) was nominally significant at Week 61 (p= 0.0428) (**Figure 1C**).

**Figure 1:**
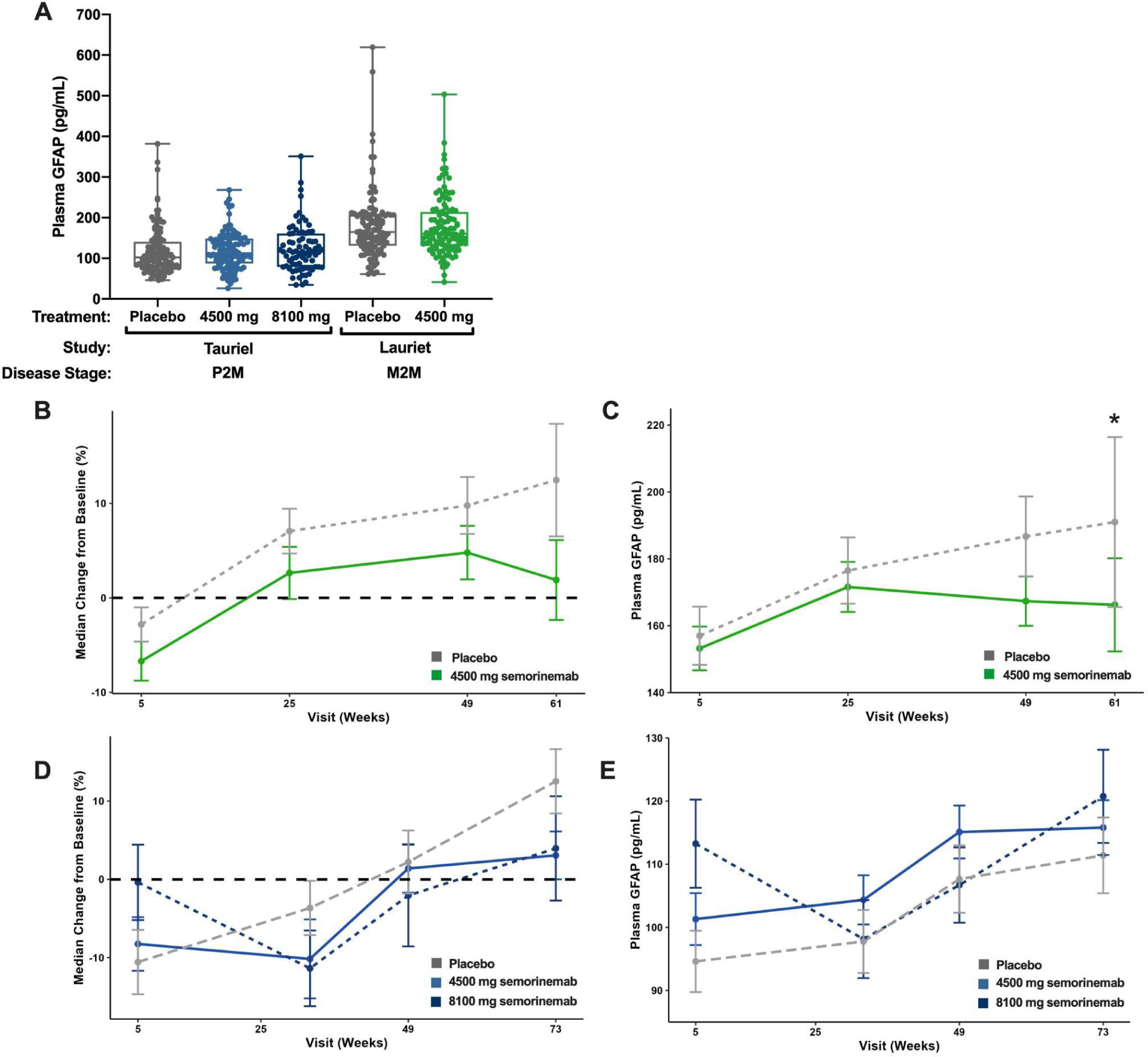
**(A)** Box plots of plasma GFAP concentrations at baseline in Tauriel and Lauriet participants across the placebo and semorinemab treatment groups **(B)** Plasma GFAP median percent change from baseline (± standard error) at Weeks 5, 25, 49, and 61 in Lauriet M2M participants treated with placebo (gray line) or 4500 mg semorinemab (green line) **(C)** Plasma GFAP median concentration (± standard error) at Weeks 5, 25, 49, and 61 in Lauriet M2M participants treated with placebo (gray line) or 4500 mg semorinemab (green line) **(D)** Plasma GFAP median percent change from baseline (± standard error) at Weeks 5, 33, 49, and 73 in Tauriel P2M participants treated with placebo (gray line), 4500 mg semorinemab (blue) and semorinemab (dark blue) **(E)** Plasma GFAP median concentration (± standard error) at Weeks 5, 33, 49, and 73 in Tauriel P2M participants treated with placebo (gray line), 4500 mg semorinemab (blue) and semorinemab (dark blue). Asterisk denotes the significance of a nominal unpaired parametric Student’s *t*-test without FDR-adjustment, where *p ≤ 0.05.

Tauriel placebo plasma GFAP levels also trended higher, but not until the end of the study period (Week 73: 12.5 ± 4.11%). In contrast to Lauriet, there was considerable overlap between the placebo and semorinemab treatment groups at Weeks 49 and 73, and no significant differences were detected between the groups irrespective of whether the evaluation was based on percent change (**Figure 1D**) or absolute concentration (**Figure 1E**). Tauriel plasma GFAP levels were not quantified from the 1500 mg semorinemab dose group.

Baseline CSF GFAP levels in the placebo and treatment groups in each trial did not differ in a statistically significant manner (**Supplemental Figure 1A**). In Tauriel, semorinemab had no effect on CSF GFAP across the dose groups. (**Supplemental Figure 1B**). Similarly, when CSF GFAP measurements from Lauriet Cohorts 1 and 2 (Weeks 49 and 61) were annualized and aggregated for analysis, concentrations were not significantly impacted by the administration of 4500 semorinemab (Weeks 49/61: 0.841 ± 2.42%) or placebo (Weeks 49/61: 2.77 ± 5.00%) (**Supplemental Figure 1C**).

To determine if there was a relationship between the plasma GFAP and cognition, an analysis of correlation between plasma GFAP concentrations and cognitive scores was performed. Baseline plasma GFAP in Tauriel participants correlated modestly, but significantly, with baseline MMSE (⍴_s_= -0.118), RBANS (⍴_s_= -0.105), and ADAS-Cog13 (⍴_s_= 0.0568). However, baseline plasma GFAP did not significantly correlate with any of the baseline cognitive and functional assessments in Lauriet (**Supplemental Table 1)**. As the 4500 mg dose of semorinemab in Lauriet was associated with a significant slowing in ADAS-Cog11, data from Lauriet Cohort 1 (Week 49) and Cohort 2 (Week 61) were annualized and then aggregated for an analysis of correlation between post-dose changes in plasma GFAP concentrations and ADAS-Cog11 scores. There was a modest correlation between reductions in plasma GFAP and ADAS-Cog11 scores in the 4500 mg semorinemab group (R_s_ = 0.235, p = 0.027) (**Figure 2A**) that was not present in the placebo group (R_s_ = 0.026, n.s) (**Figure 2B).**

**Figure 2:**
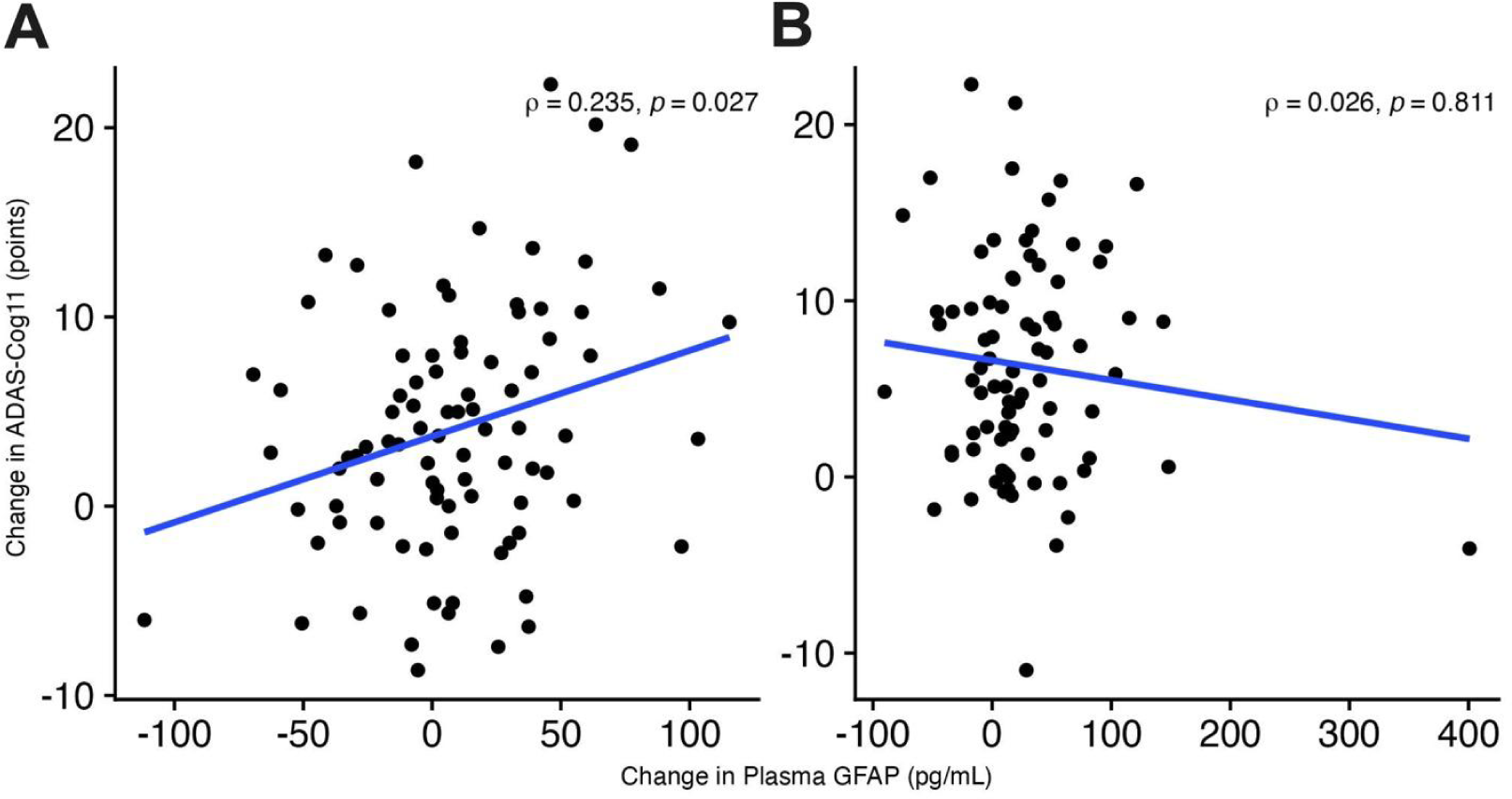
Correlations between the change from baseline at follow-up of plasma GFAP levels (pg/mL) and change from baseline at follow-up ADAS-Cog11 (points) in M2M participants treated with **(A)** 4500 mg semorinemab or **(B)** placebo. Linear regression lines fitted to the change in GFAP and ADAS-Cog11 are displayed in blue.

### YKL-40

YKL-40 levels were measured from participant CSF in Tauriel, and from participant CSF and plasma in Lauriet. Baseline CSF YKL-40 levels were statistically indistinguishable between placebo and treatment groups across trials **(Figure 3A)**. In Tauriel, CSF YKL-40 levels rose significantly after administration of semorinemab, irrespective of dose. The median percent change from baseline increased ∼29.6 - 35.2% across dose groups and timepoints. YKL-40 levels in the placebo group did not increase in this manner and were stable across the study period (∼3.2 - 4.4%). (**Figure 3B**). In Lauriet, CSF data from Cohorts 1 and 2 (Weeks 49 and 61) were annualized and then aggregated as in the CSF GFAP analysis. Consistent with Tauriel, CSF YKL-40 levels increased significantly from baseline following administration of 4500 mg semorinemab (40.7 ± 7.87%), while placebo had no effect (0.279 ± 2.66%) (**Figure 3C**). In Lauriet plasma, administration of 4500 mg semorinemab or placebo was not followed by significant changes in YKL-40 concentrations across the study period (**Supplemental Figure 2**). Plasma YKL-40 concentrations were not analyzed in Tauriel.

**Figure 3:**
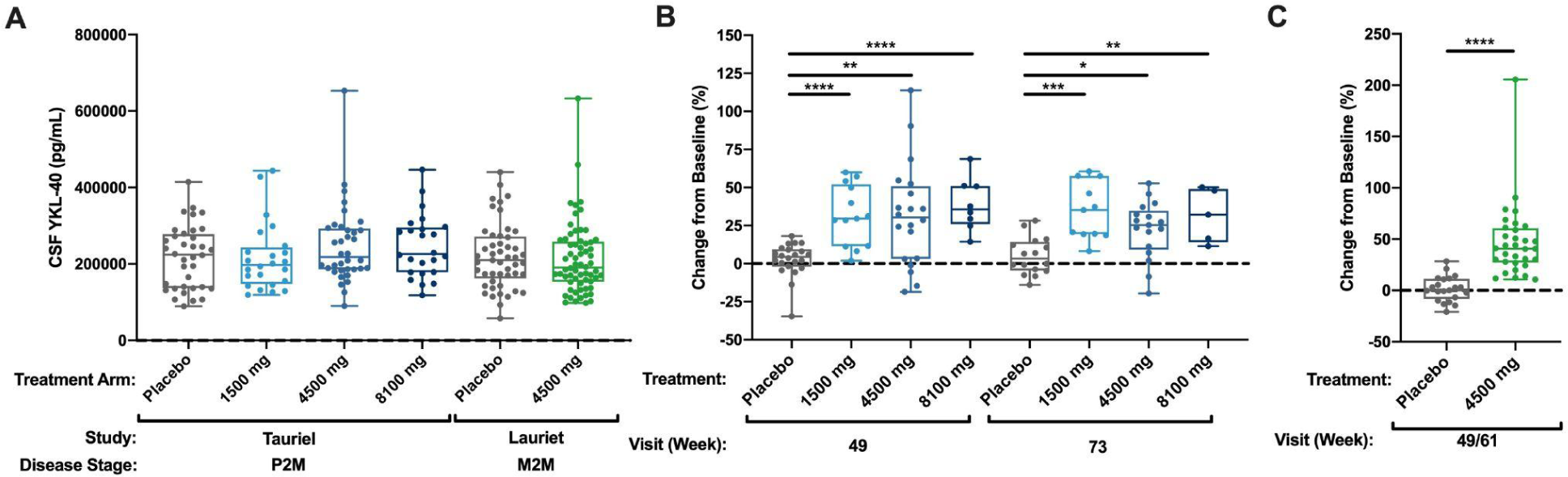
**(A)** Box plots of CSF YKL-40 concentrations at baseline in Tauriel and Lauriet participants across the placebo and semorinemab treatment groups **(B)** Boxplots of CSF YKL-40 median percent change from baseline (± standard error) at Weeks 49 and 73 in Tauriel P2M participants treated with placebo (gray), 1500 mg (light blue), 4500 mg (blue), or 8100 mg (dark blue) semorinemab. **(C)** Boxplots of annualized CSF YKL-40 median percent change from baseline (± standard error) at Weeks 49/61 in Lauriet M2M participants treated with placebo (gray line) or 4500 mg semorinemab (green). Asterisks denote the significance of an unpaired parametric Student’s *t*-test with FDR-adjustment, where ****p ≤ 0.0001, ***p ≤ 0.001, **p ≤ 0.01, *p ≤ 0.05

### Plasma pTau181

Plasma pTau181 concentrations were measured from the 4500 mg semorinemab and placebo arms of the Double Blind (DBL) treatment periods of Tauriel (Weeks 1-73) or Lauriet (Cohort 1: Weeks 1-49, Cohort 2: Weeks 1-61), and from the 4500-mg semorinemab Open Label Extension (OLE) period of Lauriet (Cohort 1: Weeks 53-145, Cohort 2: Weeks 65-157). Baseline plasma pTau181 levels showed no statistical difference between placebo and treatment groups across the trials **(Figure 4A)**. Plasma pTau181 levels increased dramatically (>30-fold over baseline) following administration of semorinemab, starting at Week 5 in both studies. (**Figure 4B)**. In Lauriet participants that received 4500 mg semorinemab in the DBL period and then continued into the OLE (23 additional Q4W doses of 4500 mg semorinemab), plasma pTau181 remained elevated for the duration of the study, demonstrating that extended exposure did not translate into reductions of this biomarker in the periphery (**Figure 4C)**. This is consistent with previously published data demonstrating that semorinemab induces a peripheral target engagement response of similar rate and magnitude when plasma total or plasma pTau217 were evaluated [1,2]. In Lauriet participants that received placebo during the DBL period and then transitioned to semorinemab in the OLE, plasma pTau181 concentrations increased to levels comparable to participants in the DBL period (>30-fold over baseline), and then remained stable for the duration of the OLE. **(Supplemental Figure 3)**.

**Figure 4:**
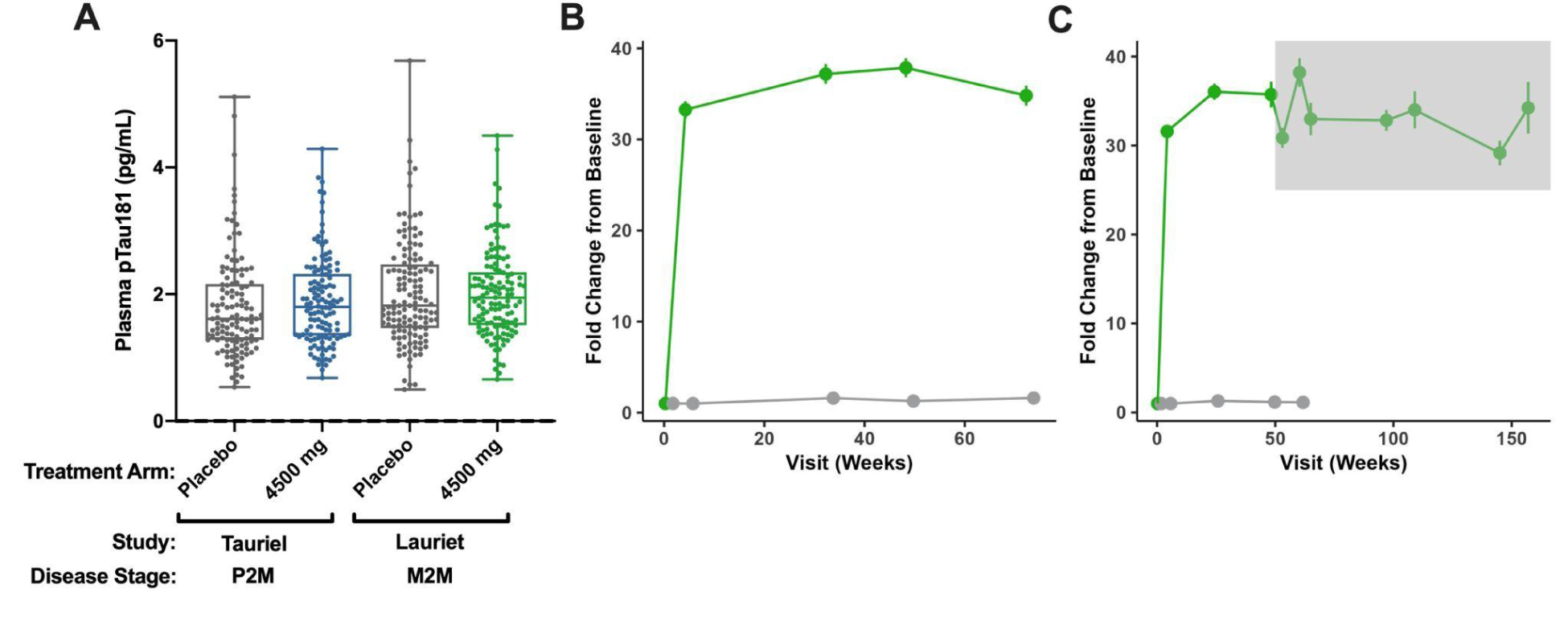
**(A)** Box plots of plasma pTau181 concentrations at baseline in Tauriel and Lauriet participants across the placebo and semorinemab treatment groups (**B**) Average fold change in plasma pTau181 concentrations over baseline in placebo (gray line) and 4500 mg semorinemab (green line) treatment groups at Weeks 1, 5, 33, 49, and 73 for Tauriel P2M participants and (**C**) at Weeks 1, 5, 25, 49, 53, 61, 65, 97, 109, 145, and 157 in Lauriet M2M participants. Measurements in the gray shaded region are from participants in OLE.

### Baseline biomarker and clinical intercorrelations

Baseline levels of AD pathophysiology biomarkers in the CSF and plasma from P2M and M2M participants enrolled in the Tauriel and Lauriet studies are shown in **Supplemental Table 2**. CSF Aβ42 concentrations were in alignment with a clinical diagnosis of AD and the presence of amyloid pathology. Concentrations were significantly lower in Lauriet participants (588 ± 17.6 pg/mL) than Tauriel participants (680 ± 15.5 pg/mL) (p <0.0001). Elevated CSF total Tau and pTau181 concentrations were also consistent with a diagnosis of AD [12]. CSF and plasma levels of NfL were significantly higher in Lauriet than Tauriel, consistent with a higher degree of neurodegeneration in the M2M population. Higher levels of CSF CXCL10, plasma GFAP, and CSF IL-6 were also present in Lauriet and consistent with more glial activity and neuroinflammation in M2M disease. Of the synaptic biomarkers, NPTX2 was significantly lower in M2M AD, consistent with reports that NPTX2 decreases with the severity of AD and predicts AD-related outcomes [13,14]. Of the remaining biomarkers, there were no significant or consistent differences between the disease populations.

We conducted an assessment of baseline correlations between plasma GFAP, CSF GFAP, and CSF YKL-40 and biomarkers shared between each study. Plasma GFAP was moderately correlated with plasma tTau (⍴_s_= 0.45), plasma pTau181 (⍴_s_= 0.53), and plasma NfL (⍴_s_= 0.54), and modestly correlated with Aβ1-42 (⍴_s_= -0.34), lipocalin (⍴_s_= -0.21), and IL-6 (⍴_s_= 0.12) (**Figure 5).** Plasma GFAP did not correlate significantly with CSF GFAP (**Supplemental Figure 4**). With the exception of IL-6 and CXCL10, CSF YKL-40 correlated moderately all CSF biomarkers examined **(Figure 5)**. As anticipated, the CSF tau species were strongly intercorrelated (⍴_s_= 0.84 - 0.97), as were plasma tTau and plasma pTau181 (⍴_s_= 0.70) **(Supplemental Figure 4)**.

**Figure 5:**
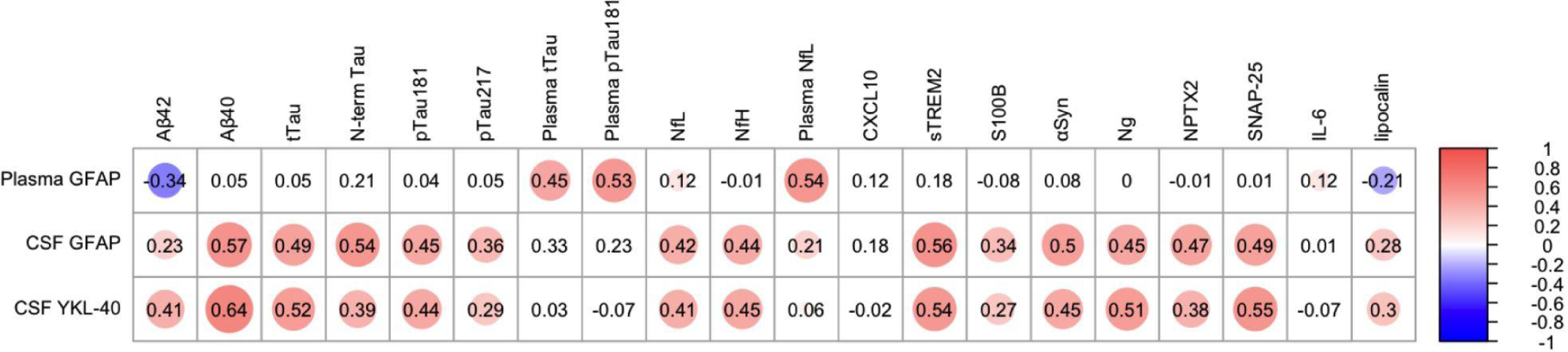
Heatmap of correlations between baseline plasma GFAP, CSF GFAP, and CSF YKL-40 levels and all other CSF and plasma biomarkers jointly evaluated in Tauriel and Lauriet. All biomarkers are measured in CSF unless specified otherwise. Spearman’s rho values are displayed and colored circles represent correlations where the FDR-adjusted p-value < 0.05.

In addition to biomarker intercorrelations, we also evaluated correlations between these analytes and cognitive/functional scores common to both studies in aggregate (CDR-SB, MMSE, ADCS-ADL). At baseline, a strong intercorrelation was observed between the shared cognitive and functional scores (⍴_s_= -0.75 - 0.51). In addition, all three cognitive/functional metrics correlated with biomarkers representative of a variety of pathophysiological features. Modest correlations were noted with plasma GFAP (⍴_s_= -0.39 - 0.29), IL-6 (⍴_s_= -0.30 - 0.25), lipocalin (⍴_s_= -0.28 - 0.34), plasma NfL (⍴_s_= -0.28 - 0.22), plasma pTau181 (⍴_s_= -0.17 - 0.18), plasma tTau (⍴_s_= -0.18 - 0.12), and CSF Aβ1-42 (⍴_s_= -0.22 - 0.37). (**Supplemental Figure 4**).

### Additional biomarkers

As outlined in Table 2, biomarkers related to neuroinflammation (**Supplemental Figure 5**), amyloidosis (**Supplemental Figure 6**), neurodegeneration (**Supplemental Figure 7**) and synaptic function (**Supplemental Figure 8**) were measured in Tauriel and Lauriet CSF and plasma. In both studies, these biomarkers did not exhibit any effects from semorinemab treatment that reached statistical significance.

## DISCUSSION

The administration of Semorinemab to P2M and M2M participants in the Tauriel and Lauriet phase 2 trials demonstrated that while target engagement could be detected in the periphery and CNS of study participants [1,2], there were divergent clinical outcomes. In a post-hoc examination of AD pathophysiology biomarkers in the plasma and CSF of trial participants, we observed noteworthy changes in glial activity biomarkers. YKL-40 increased significantly in the CSF post-dose in both studies, suggesting that the administration of semorinemab stimulated an increase in microglial activity in the CNS. The magnitude of this increase is similar to responses observed with AL002, a TREM2 agonist antibody that directly activates microglia, in a Phase 1 healthy volunteer study [15]. In the case of other drugs targeting AD pathology, no increases were reported in Crenezumab’s CREAD studies [16], and a relatively smaller increase of ∼6% was reported in Gantenerumab’s GRADUATE studies [17]. YKL-40 is expressed in microglia, astrocytes, and peripheral immune cells, and is elevated in the CSF of AD patients [18]. At the transcriptomic level, YKL-40 expression is increased in human AD brains, but not with subjects carrying the loss of function R62H TREM2 variant which reduces microglia activation [19]. Additional evidence of a semorinemab effect on microglia activity comes from proteomic analyses of Tauriel and Lauriet CSF, where the post-treatment proteomic signatures indicated that the proteomic response in Lauriet was likely of microglial origin [20]. Together with the CSF YKL-40 observations, these findings were unexpected because semorinemab was developed on an IgG4 backbone, which has been demonstrated to have lower binding affinity to fragment crystallizable gamma receptor (FcR) and expected to have reduced immune effector function [21]. However, it is possible that the binding of semorinemab to tau aggregates could drive higher local concentrations of the antibody, cross-link FcRs through an avidity effect, and trigger microglia activation.

Increases in plasma GFAP observed throughout the study periods were stabilized in response to 4500 mg semorinemab in the Lauriet trial, while no remarkable impact of treatment was observed between the dose groups in the Tauriel study. The response observed in Lauriet is in contrast to CSF GFAP, where no notable differences were observed between the placebo and treatment groups in either study. Plasma GFAP is an emerging biomarker for AD that is considered reflective of reactive astrocytes in a variety of neurodegenerative conditions [22,23]. It decreases in response to anti-amyloid therapy, is associated with plaque removal [17,24,25], and across our Ph2 studies, baseline levels demonstrate a degree of correlation with metrics of cognition and function. The robust CSF YKL-40 response and the unique enrichment of microglia proteins observed in an analysis of the Lauriet CSF proteome [20] suggest an intriguing possibility that the stabilization of plasma GFAP in Lauriet may be a response to microglia activity stimulated by semorinemab. Since semorinemab has no discernable effect on Tau aggregation, our post hoc discovery of microglia activation and variations in plasma GFAP between the trials presents a scientific hypothesis that this biology may account for the discrepancies observed in the trial outcomes. However, additional mechanistic studies need to be conducted to elucidate this hypothesis.

## Data Availability

All data produced in the present study are available upon reasonable request to the authors

---

**Supplemental Table 1:**
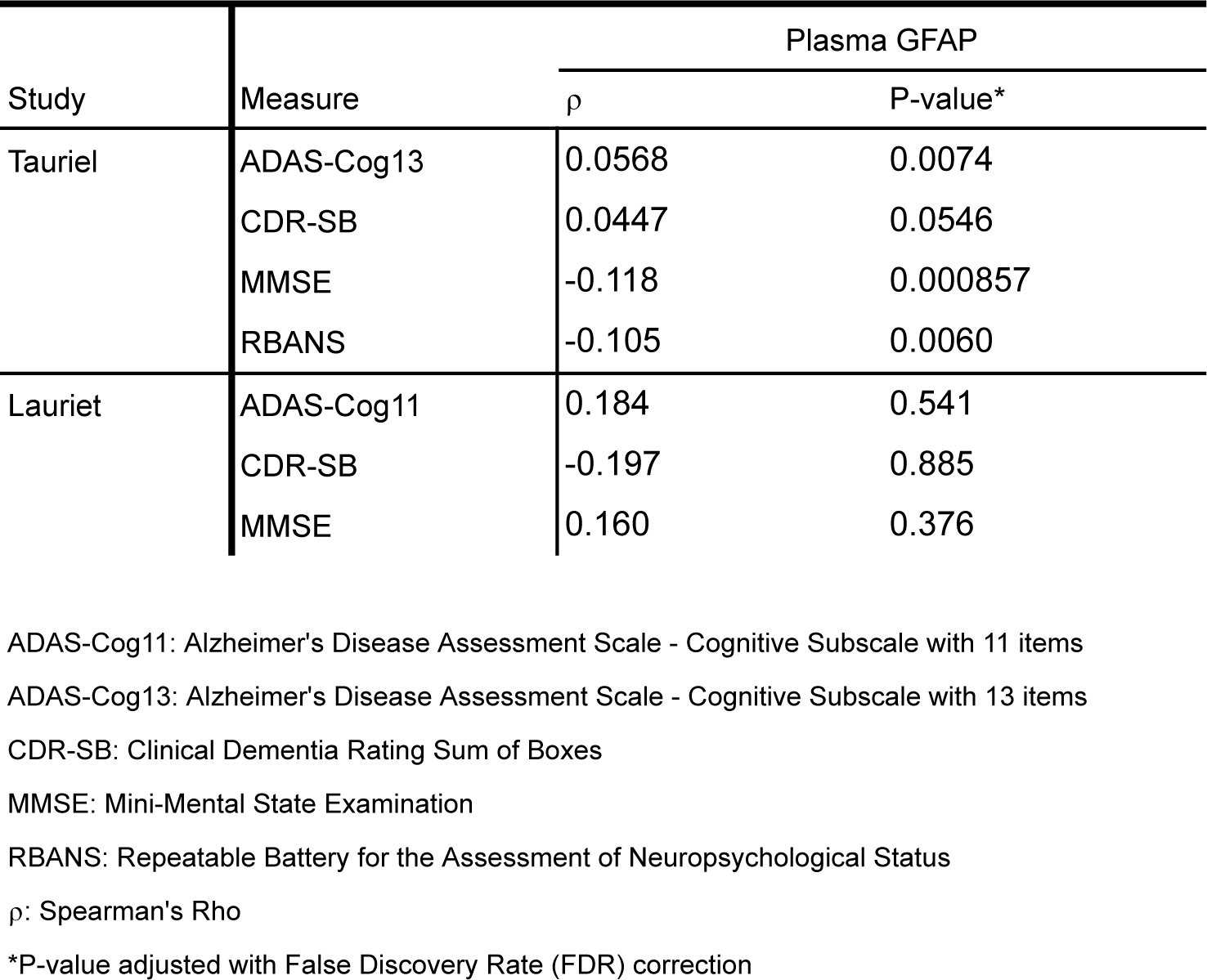
Spearman correlations between baseline plasma GFAP concentrations and cognitive metrics in Tauriel and Lauriet. P-values are adjusted using the False Discovery Rate (FDR) correction.

| Study | Measure | Plasma GFAP |  |
| --- | --- | --- | --- |
| | | $\rho$ | P-value* |
| Tauriel | ADAS-Cog13 | 0.0568 | 0.0074 |
|  | CDR-SB | 0.0447 | 0.0546 |
|  | MMSE | -0.118 | 0.000857 |
|  | RBANS | -0.105 | 0.0060 |
| Lauriet | ADAS-Cog11 | 0.184 | 0.541 |
|  | CDR-SB | -0.197 | 0.885 |
|  | MMSE | 0.160 | 0.376 |

ADAS-Cog11: Alzheimer's Disease Assessment Scale - Cognitive Subscale with 11 items
ADAS-Cog13: Alzheimer's Disease Assessment Scale - Cognitive Subscale with 13 items
CDR-SB: Clinical Dementia Rating Sum of Boxes
MMSE: Mini-Mental State Examination
RBANS: Repeatable Battery for the Assessment of Neuropsychological Status
$\rho$ : Spearman's Rho
\*P-value adjusted with False Discovery Rate (FDR) correction

**Supplemental Table 2:**
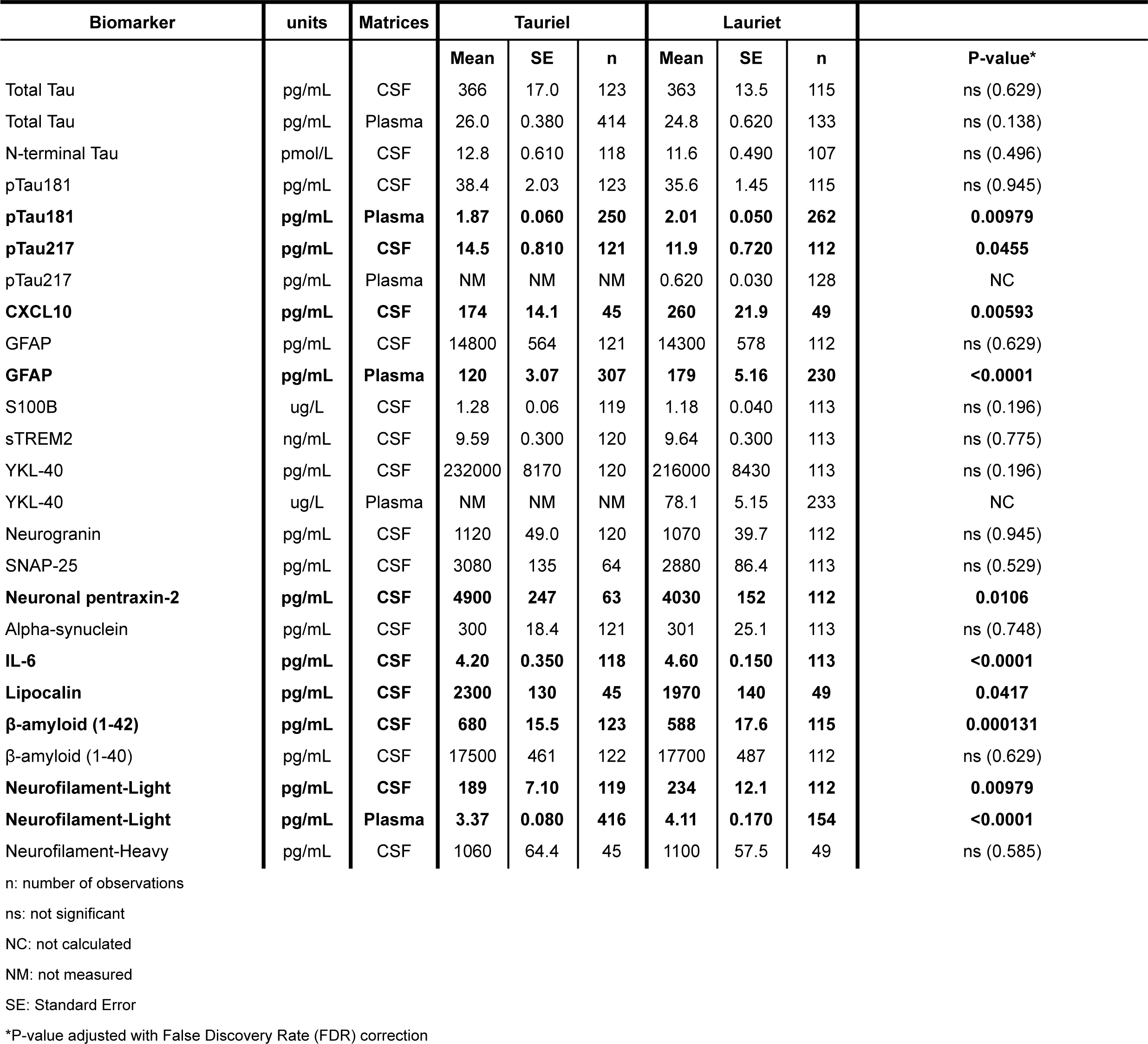
Mean +/- standard error of biomarker concentrations at baseline and between studies. Mann-Whitney p-values are adjusted using the False Discovery Rate (FDR) correction.

**Supplemental Figure 1:**
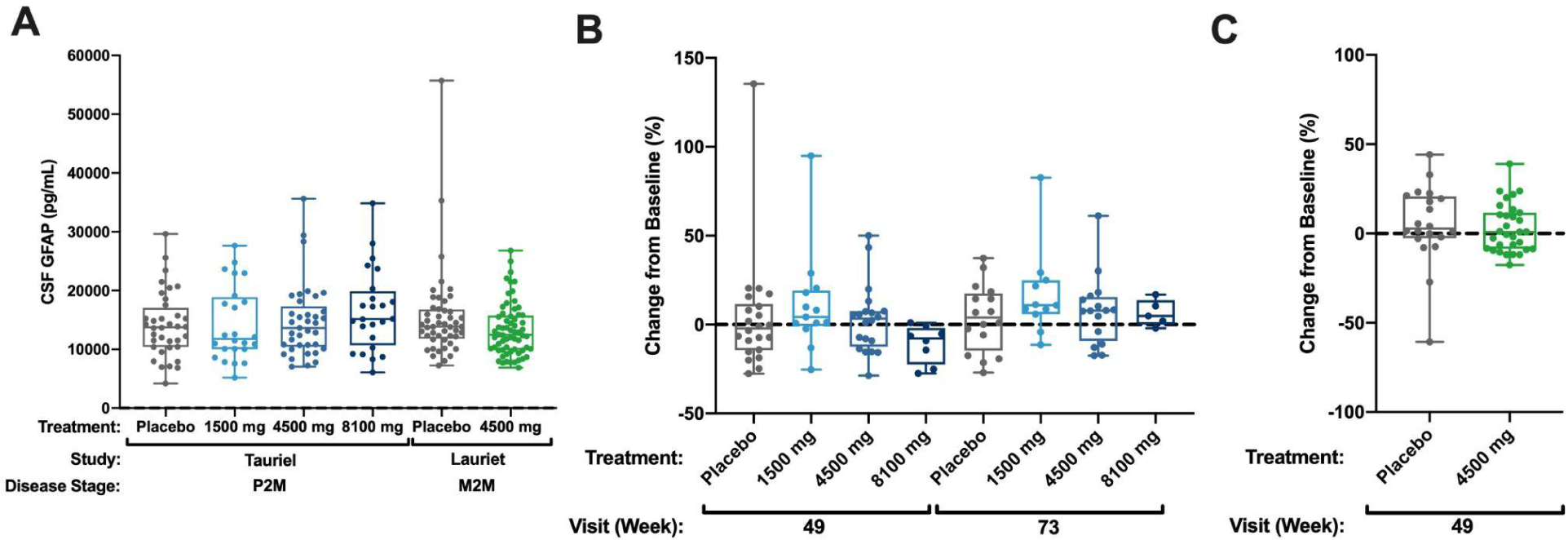
**(A)** Boxplots of CSF GFAP concentrations at baseline in Tauriel and Lauriet participants across the placebo and semorinemab treatment groups **(B)** Boxplots of CSF GFAP median percent change from baseline (± standard error) at Weeks 49 and 73 in Tauriel P2M participants treated with placebo (gray), 1500 mg (light blue), 4500 mg (blue), or 8100 mg (dark blue) semorinemab. **(C)** Boxplots of annualized CSF GFAP median percent change from baseline (± standard error) at Weeks 49/61 in Lauriet M2M participants treated with placebo (gray line) or 4500 mg semorinemab (green).

**Supplemental Figure 2:**
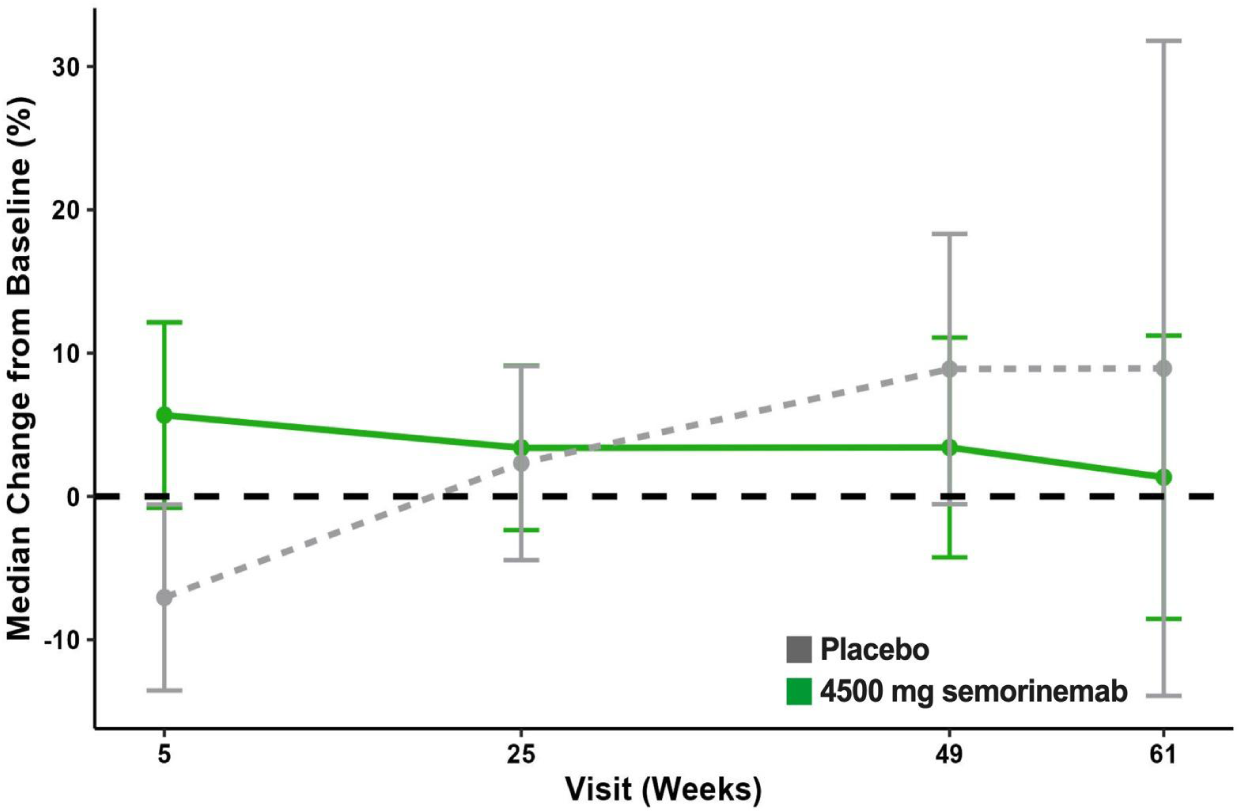
Plasma YKL-40 median concentration (± standard error) at Weeks 5, 25, 49, and 61 in Lauriet M2M participants treated with placebo (gray line) or 4500 mg semorinemab (green line).

**Supplemental Figure 3:**
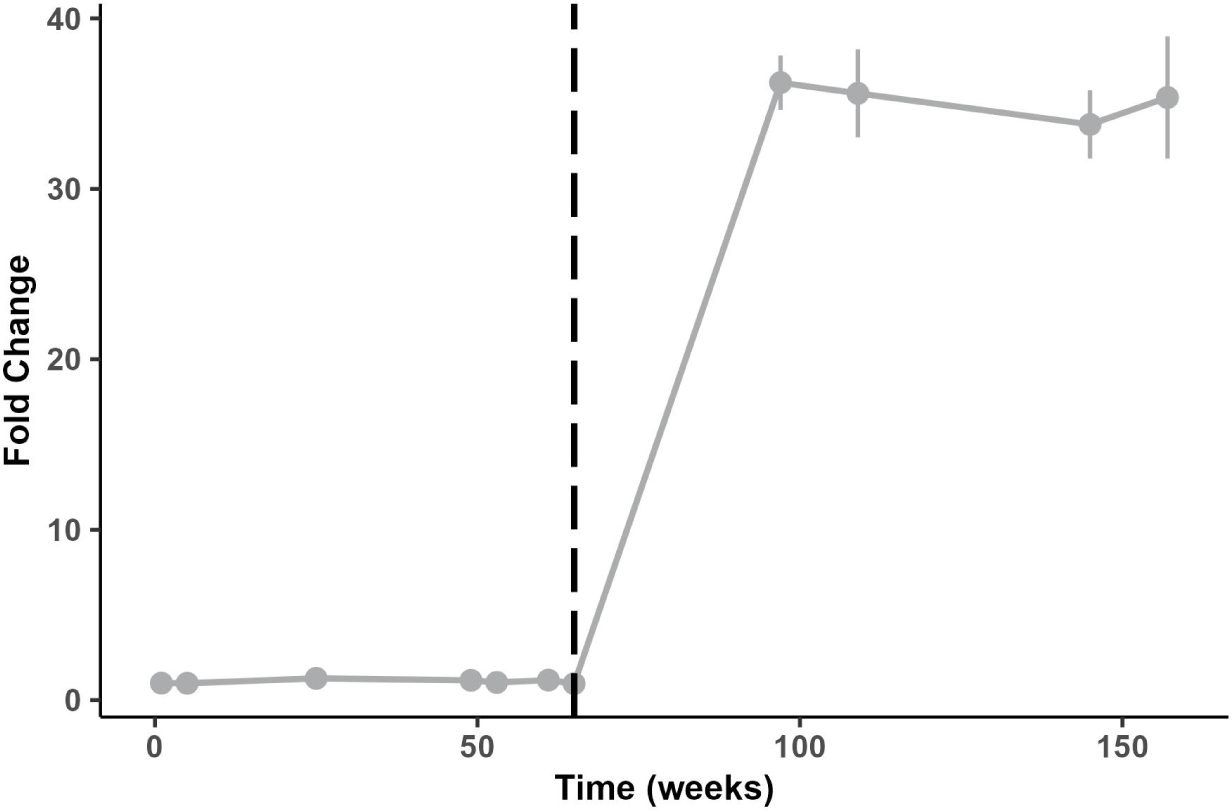
Average fold change of plasma pTau181 concentrations over baseline at Weeks 1, 5, 25, 49, 53, 61, 65, 97, 109, 145, and 157 in Lauriet M2M participants who transitioned from placebo to 4500 mg semorinemab in the OLE. Dashed line denotes conversion from placebo to active treatment.

**Supplemental Figure 4:**
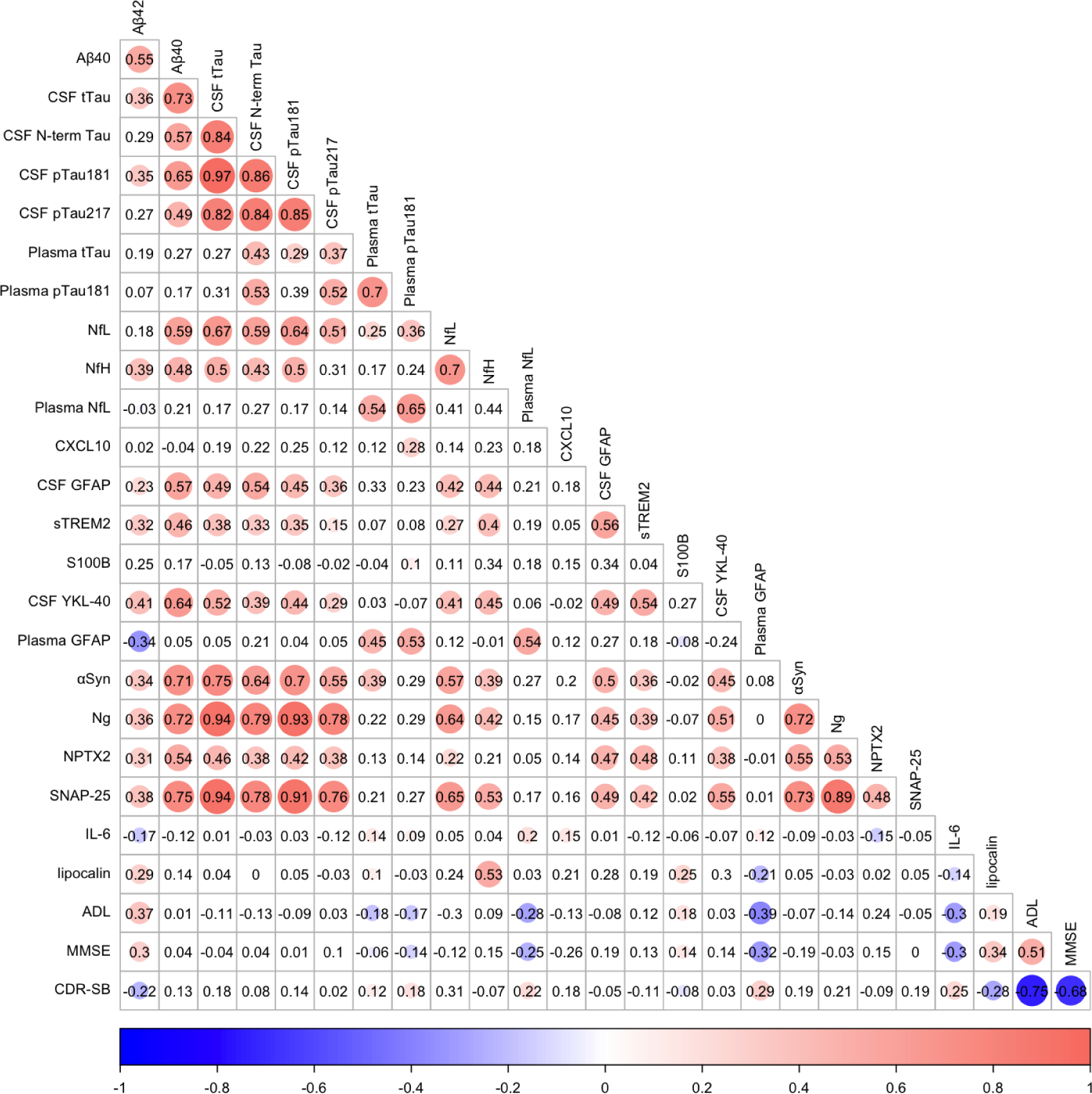
Heatmap of correlations between baseline biomarker concentrations and clinical measures jointly evaluated in Tauriel and Lauriet. All biomarkers are measured in CSF unless specified otherwise. Spearman’s rho values are displayed and colored circles represent correlations where the FDR-adjusted p-value < 0.05.

**Supplemental Figure 5:**
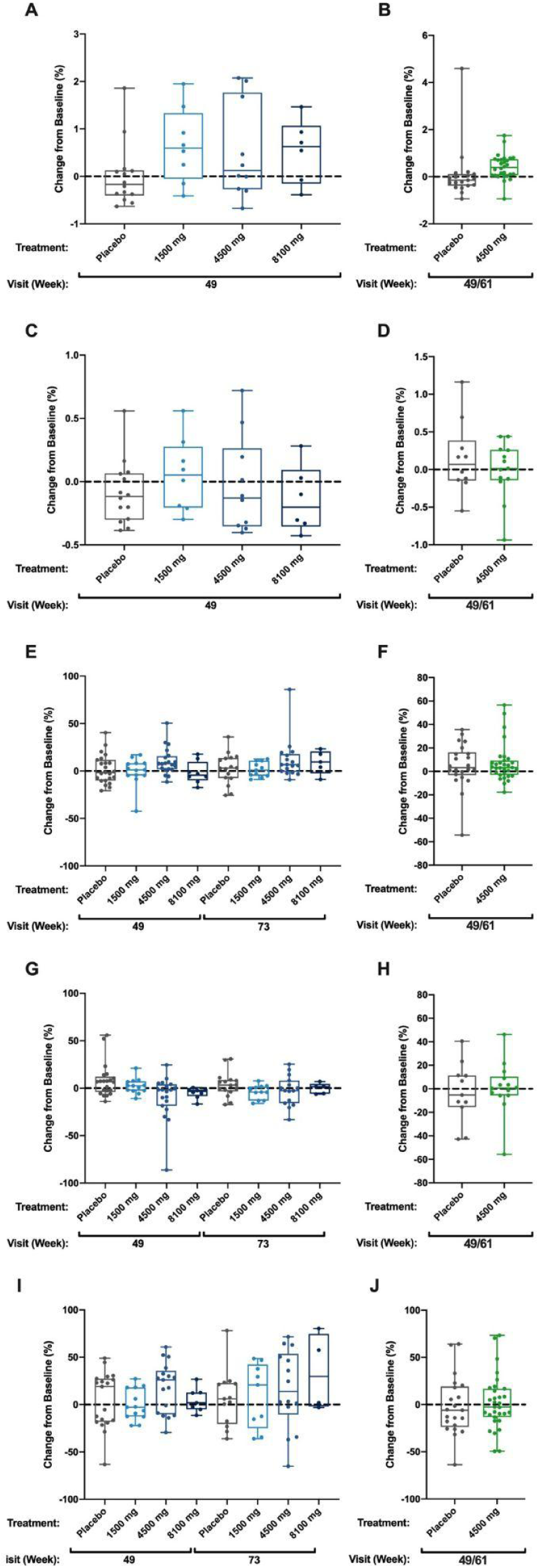
CSF neuroinflammation biomarkers **(A)** Boxplots of CSF CXCL10 median percent change from baseline (± standard error) at Week 49 in Tauriel P2M participants treated with placebo (gray), 1500 mg (light blue), 4500 mg (blue), or 8100 mg (dark blue) semorinemab. **(B)** Boxplots of annualized CSF CXCL10 median percent change from baseline (± standard error) at Weeks 49/61 in Lauriet M2M participants treated with placebo (gray) or 4500 mg semorinemab (green). **(C)** Box plots of CSF lipocalin median percent change from baseline (± standard error) at Week 49 in Tauriel P2M participants treated with placebo (gray), 1500 mg (light blue), 4500 mg (blue), or 8100 mg (dark blue) semorinemab. **(D)** Boxplots of annualized CSF lipocalin median percent change from baseline (± standard error) at Weeks 49/61 in Lauriet M2M participants treated with placebo (gray) or 4500 mg semorinemab (green). **(E)** Boxplots of CSF sTREM2 median percent change from baseline (± standard error) at Weeks 49 and 73 in Tauriel P2M participants treated with placebo (gray), 1500 mg (light blue), 4500 mg (blue), or 8100 mg (dark blue) semorinemab. **(F)** Boxplots of annualized CSF sTREM2 median percent change from baseline (± standard error) at Weeks 49/61 in Lauriet M2M participants treated with placebo (gray) or 4500 mg semorinemab (green). **(G)** Boxplots of CSF S100B median percent change from baseline (± standard error) at Weeks 49 and 73 in Tauriel P2M participants treated with placebo (gray), 1500 mg (light blue), 4500 mg (blue), or 8100 mg (dark blue) semorinemab. **(H)** Boxplots of annualized CSF S100B median percent change from baseline (± standard error) at Weeks 49/61 in Lauriet M2M participants treated with placebo (gray) or 4500 mg semorinemab (green). **(I)** Boxplots of CSF IL-6 median percent change from baseline (± standard error) at Weeks 49 and 73 in Tauriel P2M participants treated with placebo (gray), 1500 mg (light blue), 4500 mg (blue), or 8100 mg (dark blue) semorinemab. **(J)** Boxplots of annualized CSF IL-6 median percent change from baseline (± standard error) at Weeks 49/61 in Lauriet M2M participants treated with placebo (gray) or 4500 mg semorinemab (green).

**Supplemental Figure 6:**
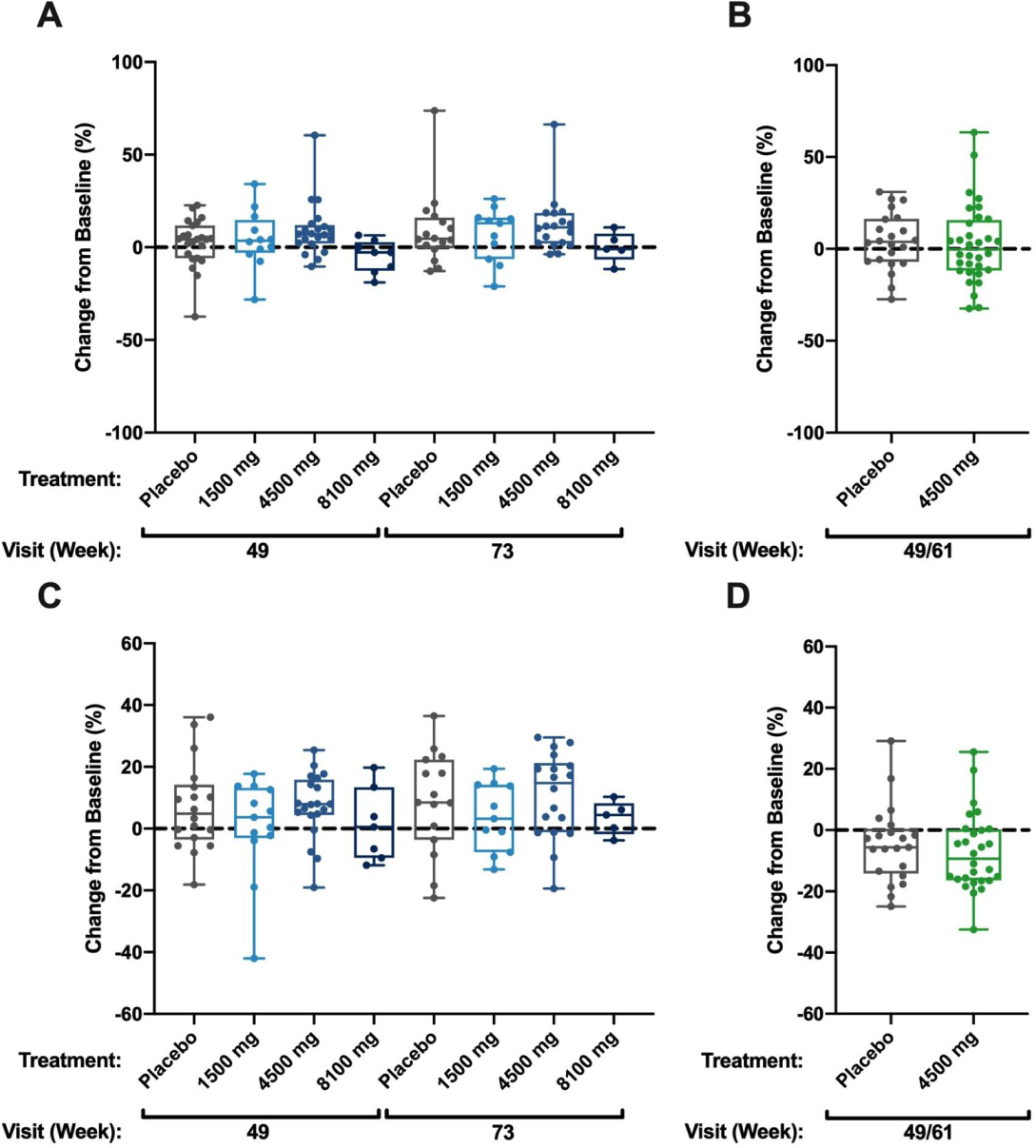
CSF amyloidosis biomarkers **(A)** Boxplots of CSF Aβ1-42 median percent change from baseline (± standard error) at Weeks 49 and 73 in Tauriel P2M participants treated with placebo (gray), 1500 mg (light blue), 4500 mg (blue), or 8100 mg (dark blue) semorinemab. **(B)** Boxplots of annualized CSF Aβ1-42 median percent change from baseline (± standard error) at Weeks 49/61 in Lauriet M2M participants treated with placebo (gray) or 4500 mg semorinemab (green). **(C)** Box plots of CSF Aβ1-40 median percent change from baseline (± standard error) at Weeks 49 and 73 in Tauriel P2M participants treated with placebo (gray), 1500 mg (light blue), 4500 mg (blue), or 8100 mg (dark blue) semorinemab. **(D)** Boxplots of annualized CSF Aβ1-40 median percent change from baseline (± standard error) at Weeks 49/61 in Lauriet M2M participants treated with placebo (gray) or 4500 mg semorinemab (green).

**Supplemental Figure 7:**
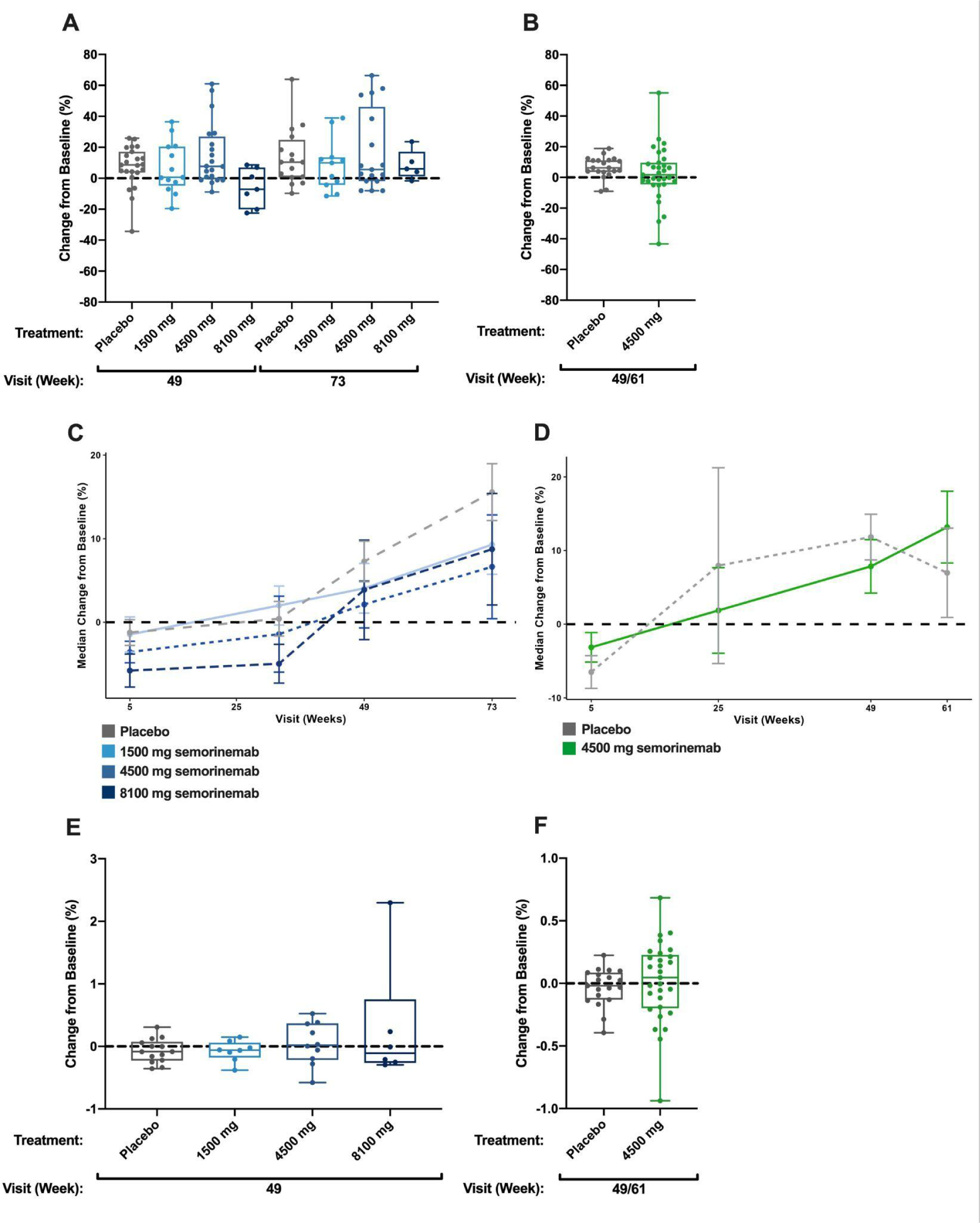
Neurodegeneration biomarkers **(A)** Boxplots of CSF NfL median percent change from baseline (± standard error) at Weeks 49 and 73 in Tauriel P2M participants treated with placebo (gray), 1500 mg (light blue), 4500 mg (blue), or 8100 mg (dark blue) semorinemab. **(B)** Boxplots of annualized CSF NfL median percent change from baseline (± standard error) at Weeks 49/61 in Lauriet M2M participants treated with placebo (gray) or 4500 mg semorinemab (green) **(C)** Plasma NfL median percent change from baseline (± standard error) at Weeks 5, 33, 49, and 73 in Tauriel P2M participants treated with placebo (gray line), 1500 mg (light blue line), 4500 mg (blue line), or 8100 mg (dark blue line) semorinemab **(D)** Plasma NfL median percent change from baseline (± standard error) at Weeks 5, 25, 49, and 61 in Lauriet M2M participants treated with placebo (gray line) or 4500 mg semorinemab (green line) **(E)** Box plots of CSF NfH median percent change from baseline (± standard error) at Week 49 in Tauriel P2M participants treated with placebo (gray), 1500 mg (light blue), 4500 mg (blue), or 8100 mg (dark blue) semorinemab. **(F)** Boxplots of annualized CSF NfH median percent change from baseline (± standard error) at Weeks 49/61 in Lauriet M2M participants treated with placebo (gray) or 4500 mg semorinemab (green).

**Supplemental Figure 8:**
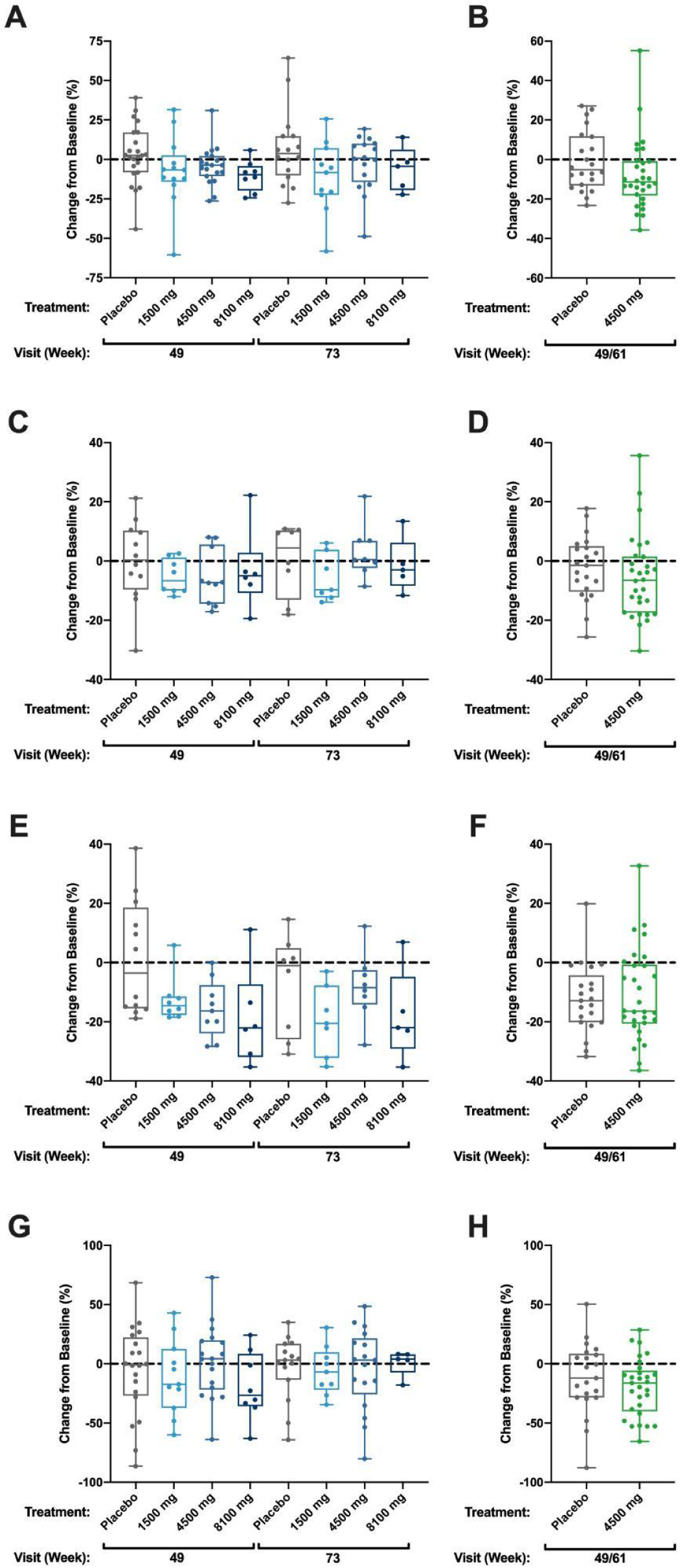
Synaptic biomarkers **(A)** Box plots of CSF Ng median percent change from baseline (± standard error) at Weeks 49 and 73 in Tauriel P2M participants treated with placebo (gray), 1500 mg (light blue), 4500 mg (blue), or 8100 mg (dark blue) semorinemab. **(B)** Boxplots of annualized CSF Ng median percent change from baseline (± standard error) at Weeks 49/61 in Lauriet M2M participants treated with placebo (gray) or 4500 mg semorinemab (green). **(C)** Box plots of CSF SNAP-25 median percent change from baseline (± standard error) at Weeks 49 and 73 in Tauriel P2M participants treated with placebo (gray), 1500 mg (light blue), 4500 mg (blue), or 8100 mg (dark blue) semorinemab. **(D)** Boxplots of annualized CSF SNAP-25 median percent change from baseline (± standard error) at Weeks 49/61 in Lauriet M2M participants treated with placebo (gray) or 4500 mg semorinemab (green). **(E)** Box plots of CSF NPTX2 median percent change from baseline (± standard error) at Weeks 49 and 73 in Tauriel P2M participants treated with placebo (gray), 1500 mg (light blue), 4500 mg (blue), or 8100 mg (dark blue) semorinemab. **(F)** Boxplots of annualized CSF NPTX2 median percent change from baseline (± standard error) at Weeks 49/61 in Lauriet M2M participants treated with placebo (gray) or 4500 mg semorinemab (green). **(G)** Box plots of CSF ɑSyn median percent change from baseline (± standard error) at Weeks 49 and 73 in Tauriel P2M participants treated with placebo (gray), 1500 mg (light blue), 4500 mg (blue), or 8100 mg (dark blue) semorinemab. **(H)** Boxplots of annualized CSF ɑSyn median percent change from baseline (± standard error) at Weeks 49/61 in Lauriet M2M participants treated with placebo (gray) or 4500 mg semorinemab (green).

## Acknowledgements/Conflicts/Funding Sources

ET, CM, KW: Study design. AB, GK, JL, JJ, NW: Sample and data acquisition. BT and SS: Data analysis. BT, CM, ET, FY, LH, VA, SS: Data interpretation. SS, ET, FY: Manuscript writing - original draft. All authors: Manuscript writing - review and editing. We would like to extend our thanks to Sarah Magazu (Roche Diagnostics GmbH) for her contributions to the development of the Elecsys plasma pTau181 extended range RPA.

The NeuroToolKit is a panel of exploratory prototype assays designed to robustly evaluate biomarkers associated with key pathologic events characteristic of AD and other neurological disorders, used for research purposes only and not approved for clinical use (Roche Diagnostics International Ltd, Rotkreuz, Switzerland). Elecsys β-Amyloid (1-42), Total-Tau, and Phospho-Tau (181P) CSF assays are approved for clinical use.

COBAS and ELECSYS are trademarks of Roche. All other product names and trademarks are the property of their respective owners.

AB, GK, and NW are full-time employees of Roche Diagnostics GmbH, Penzberg, Germany. All other authors are full-time employees of Genentech Inc. (member of the oche group). Edmond Teng is listed as a co-inventor on the patent for semorinemab.

## REFERENCES

[1] Teng E, Manser PT, Pickthorn K, Brunstein F, Blendstrup M, Bohorquez SS, et al. Safety and Efficacy of Semorinemab in Individuals With Prodromal to Mild Alzheimer Disease. Jama Neurol 2022;79:758–67. 10.1001/jamaneurol.2022.1375.

[2] Monteiro C, Toth B, Brunstein F, Bobbala A, Datta S, Ceniceros R, et al. A Randomized Phase II Study of the Safety and Efficacy of Semorinemab in Participants with Mild-to-Moderate Alzheimer’s Disease (Lauriet). Neurology 2023:10.1212/WNL.0000000000207663. 10.1212/wnl.0000000000207663.

[3] 2023 Alzheimer’s disease facts and figures. Alzheimer’s Dement 2023;19:1598–695. 10.1002/alz.13016.

[4] Knopman DS, Amieva H, Petersen RC, Chételat G, Holtzman DM, Hyman BT, et al. Alzheimer disease. Nat Rev Dis Primers 2021;7:33. 10.1038/s41572-021-00269-y.

[5] Braak H, Braak E. Neuropathological stageing of Alzheimer-related changes. Acta Neuropathol 1991;82:239–59. 10.1007/bf00308809.

[6] Braak H, Braak E. Staging of alzheimer’s disease-related neurofibrillary changes. Neurobiol Aging 1995;16:271–8. 10.1016/0197-4580(95)00021-6.

[7] Giannakopoulos P, Herrmann FR, Bussière T, Bouras C, Kövari E, Perl DP, et al. Tangle and neuron numbers, but not amyloid load, predict cognitive status in Alzheimer’s disease. Neurology 2003;60:1495–500. 10.1212/01.wnl.0000063311.58879.01.

[8] Pontecorvo MJ, Devous MD, Navitsky M, Lu M, Salloway S, Schaerf FW, et al. Relationships between flortaucipir PET tau binding and amyloid burden, clinical diagnosis, age and cognition. Brain 2017;140:748–63. 10.1093/brain/aww334.

[9] Bloom GS. Amyloid-β and Tau: The Trigger and Bullet in Alzheimer Disease Pathogenesis. JAMA Neurol 2014;71:505–8. 10.1001/jamaneurol.2013.5847.

[10] Braak H, Alafuzoff I, Arzberger T, Kretzschmar H, Tredici KD. Staging of Alzheimer disease-associated neurofibrillary pathology using paraffin sections and immunocytochemistry. Acta Neuropathol 2006;112:389–404. 10.1007/s00401-006-0127-z.

[11] Dujardin S, Hyman BT. Tau Prion-Like Propagation: State of the Art and Current Challenges. Adv Exp Med Biol 2019;1184:305–25. 10.1007/978-981-32-9358-8_23.

[12] Willemse EAJ, Maurik IS van, Tijms BM, Bouwman FH, Franke A, Hubeek I, et al. Diagnostic performance of Elecsys immunoassays for cerebrospinal fluid Alzheimer’s disease biomarkers in a nonacademic, multicenter memory clinic cohort: The ABIDE project. Alzheimer’s Dement: Diagn, Assess Dis Monit 2018;10:563–72. 10.1016/j.dadm.2018.08.006.

[13] Begcevic I, Tsolaki M, Brinc D, Brown M, Martinez-Morillo E, Lazarou I, et al. Neuronal pentraxin receptor-1 is a new cerebrospinal fluid biomarker of Alzheimer’s disease progression. F1000Research 2018;7:1012. 10.12688/f1000research.15095.1.

[14] Swanson A, Willette AA, Initiative for the ADN. Neuronal Pentraxin 2 predicts medial temporal atrophy and memory decline across the Alzheimer’s disease spectrum. Brain, Behav, Immun 2016;58:201–8. 10.1016/j.bbi.2016.07.148.

[15] Ward M, Long H, Schwabe T, Rhinn H, Tassi I, Salazar SV, et al. A phase 1 study of AL002 in healthy volunteers. Alzheimer’s Dement 2021;17. 10.1002/alz.054669.

[16] Ostrowitzki S, Bittner T, Sink KM, Mackey H, Rabe C, Honig LS, et al. Evaluating the Safety and Efficacy of Crenezumab vs Placebo in Adults With Early Alzheimer Disease. JAMA Neurol 2022;79:1113–21. 10.1001/jamaneurol.2022.2909.

[17] Bittner T, Blennow K, Scelsi M, Palermo G, Kollmorgen G, Smith J, et al. ADPD-2023-presentation-bittner-graduate-I-and-II-results-effect-of-subcutaneous.pdf. Medically 2023.

[18] Llorens F, Thüne K, Tahir W, Kanata E, Diaz-Lucena D, Xanthopoulos K, et al. YKL-40 in the brain and cerebrospinal fluid of neurodegenerative dementias. Mol Neurodegener 2017;12:83. 10.1186/s13024-017-0226-4.

[19] Zhou Y, Song WM, Andhey PS, Swain A, Levy T, Miller KR, et al. Human and mouse single-nucleus transcriptomics reveal TREM2-dependent and TREM2-independent cellular responses in Alzheimer’s disease. Nat Med 2020;26:131–42. 10.1038/s41591-019-0695-9.

[20] Abdel-Haleem AM, Casavant E, Toth B, Teng E, Monteiro C, Pandya NJ, et al. CSF proteomic analysis of semorinemab Ph2 trials in prodromal-to-mild (Tauriel) and mild-to-moderate (Lauriet) Alzheimer’s disease identifies distinct trial cell-type specific proteomic signatures. MedRxiv 2024:2024.04.11.24305670. 10.1101/2024.04.11.24305670.

[21] Ayalon G, Lee S-H, Adolfsson O, Foo-Atkins C, Atwal JK, Blendstrup M, et al. Antibody semorinemab reduces tau pathology in a transgenic mouse model and engages tau in patients with Alzheimer’s disease. Sci Transl Med 2021;13. 10.1126/scitranslmed.abb2639.

[22] Abdelhak A, Foschi M, Abu-Rumeileh S, Yue JK, D’Anna L, Huss A, et al. Blood GFAP as an emerging biomarker in brain and spinal cord disorders. Nat Rev Neurol 2022;18:158–72. 10.1038/s41582-021-00616-3.

[23] Liddelow SA, Barres BA. Reactive Astrocytes: Production, Function, and Therapeutic Potential. Immunity 2017;46:957–67. 10.1016/j.immuni.2017.06.006.

[24] Dyck CH van, Swanson CJ, Aisen P, Bateman RJ, Chen C, Gee M, et al. Lecanemab in Early Alzheimer’s Disease. New Engl J Med 2022;388:9–21. 10.1056/nejmoa2212948.

[25] Pontecorvo MJ, Lu M, Burnham SC, Schade AE, Dage JL, Shcherbinin S, et al. Association of Donanemab Treatment With Exploratory Plasma Biomarkers in Early Symptomatic Alzheimer Disease. Jama Neurol 2022;79:1250–9. 10.1001/jamaneurol.2022.3392.

